# Barriers and Facilitators to Trustworthy and Ethical AI-enabled Medical Care From Patient’s and Healthcare Provider’s Perspectives: A Literature Review

**DOI:** 10.1101/2023.10.02.23296447

**Authors:** Maryam Mooghali, Austin M. Stroud, Dong Whi Yoo, Barbara A Barry, Alyssa A. Grimshaw, Joseph S. Ross, Xuan Zhu, Jennifer E. Miller

## Abstract

**Background:** Artificial intelligence (AI) and machine learning (ML) are increasingly used for prevention, diagnosis, monitoring, and treatment of cardiovascular diseases. Despite the potential for AI/ML to improve care, ethical concerns and mistrust in AI-enabled health care exist among the public and medical community. To inform practice guidelines and regulatory policies that facilitate ethical and trustworthy use of AI in medicine, we conducted a literature review to identify key ethical and trust barriers and facilitators from patients’ and healthcare providers’ perspectives when using AI in cardiovascular care.

**Methods:** In this rapid literature review, we searched six bibliographic databases to identify publications discussing transparency, trust, or ethical concerns (outcomes of interest) associated with AI/ML-based medical devices (interventions of interest) in the context of cardiovascular care from patients’, caregivers’, or healthcare providers’ perspectives. The search was completed on May 24, 2022 and was not limited by date or study design.

**Results:** After reviewing 7,925 papers from six databases and 3,603 papers identified through citation chasing, 145 articles were included. Key ethical concerns included privacy, security, or confidentiality issues; risk of healthcare inequity or disparity; risk of patient harm; accountability and responsibility concerns; problematic informed consent and potential loss of patient autonomy; and issues related to data ownership. Major trust barriers included data privacy and security concerns, potential risk of patient harm, perceived lack of transparency about AI-enabled medical devices, concerns about AI replacing human aspects of care, concerns about prioritizing profits over patients’ interests, and lack of robust evidence related to the accuracy and limitations of AI-based medical devices. Ethical and trust facilitators included ensuring data privacy and data validation, conducting clinical trials in diverse cohorts, providing appropriate training and resources to patients and healthcare providers and improving their engagement in different phases of AI implementation, and establishing further regulatory oversights.

**Conclusion:** This review revealed key ethical concerns and barriers and facilitators of trust in AI-enabled medical devices from patients’ and healthcare providers’ perspectives. Mitigation strategies, including enhancing regulatory oversight on the use of patient data and promoting AI safety and transparency are needed for effective implementation of AI in cardiovascular care.

## BACKGROUND

Artificial intelligence (AI) and machine learning (ML) are increasingly used in healthcare to improve the prevention, diagnosis, treatment, and maintenance of health conditions.^1^ These interventions have enormous potential to assist in the management of cardiovascular diseases, the leading cause of death in the US, given the high number of AI-based devices authorized for use and under review by the FDA for cardiovascular diseases, the breadth of use cases spanning clinical practice to consumer-facing AI-enabled solutions, and the potential for improving clinical outcomes.^2–5^

Previous studies have shown that patients may be willing to accept the use of AI in healthcare and see its potential benefits if certain conditions are met, including transparency about the capture and use of their data by AI systems and the ability to opt out from data sharing at any time.^6^ Moreover, patients place a higher level of trust in a healthcare provider’s assessment of their health compared to an AI and often want assurance that their physicians are involved in and ultimately are responsible for AI-enabled decisions due to the concerns about risks of AI failures during care.^7,8^ On a similar note, healthcare providers express specific needs for information transparency, such as explanations about known strengths and limitations of interventions when using AI-based software in clinical decision-making.^9^ Healthcare providers also recognize the potential impact of AI on patient-clinician trust and seek support for transparent and effective communication with patients about AI use in their care.^10^ Thus, to fully achieve the appropriate uptake of AI/ML in medicine, patients’ and healthcare providers’ ethical and trust concerns must be addressed.^11^

Although existing research has begun to explore general patient and clinician perspectives on AI transparency, trust, and ethical concerns, specific barriers and facilitators to addressing these stakeholder concerns when implementing AI in cardiovascular care remain sparse and largely unactionable. More systematic research is needed to uncover the nuanced requirements for trusted understanding and use of these AI/ML-based medical devices and to inform practice guidelines and regulatory policies that facilitate ethical and trustworthy use of AI in medicine. Accordingly, this study reviews the literature to identify key ethical concerns, potential mitigation strategies, and barriers and facilitators to trustworthy AI-informed cardiovascular care.

## METHODS

### Inclusion and Exclusion Criteria

We conducted a rapid review of the literature, a form of information synthesis aiming to generate evidence through a resource-efficient approach by simplifying or removing certain components of the traditional systematic review process.^12^ Eligible for inclusion were publications discussing transparency, trust, or ethical concerns (outcomes of interest) associated with AI/ML-based medical devices (interventions of interest) in the context of cardiovascular care from patients’, caregivers’, or healthcare providers’ perspectives. Our search was not limited by date or study design. All papers published as full manuscripts, including qualitative and quantitative analyses, commentaries, editorials, expert opinions, perspective pieces, and guidelines were included. Conference abstracts, book chapters, pre-prints, animal studies, and publications that were not in English were excluded. Prior to the formal article screening process, a calibration exercise was conducted to test our screening criteria.

### Search Strategy and Data Sources

A medical librarian with literature review expertise (AAG) developed the search strategy with input from all authors. The search was developed as an Ovid Embase search strategy, which was subsequently reviewed by a second librarian not otherwise associated with the project using PRESS.^13^ After the strategy had been finalized and unanimously approved by all authors, it was adapted to the syntax and subject headings of other databases. Details on the search strategy can be found in Error! Reference source not found.. The search was conducted on the following six bibliographic databases: Cochrane Library, Embase, Google Scholar, Ovid Medline, Scopus, and Web of Science Core Collection, and was completed on May 24, 2022.

### Study Selection

Search results were downloaded to EndNote 20 (Clarivate, Philadelphia, PA), and duplicate citations were removed using the Yale Deduplicator Tool.^14^ Individual citations were ingested into Covidence, a software tool dedicated to literature review management that facilitates collaboration between independent reviewers in the article screening and review processes. The review process was divided into two major steps: title/abstract screening and full-text screening. Titles and abstracts of each paper identified by the search were independently screened by two authors [MM and AMS, AAG, or DWY] against the inclusion criteria. Next, full-text articles were obtained for all studies that had not been excluded at the first level of screening and were assessed by two independent reviewers [MM and AMS or DWY], with the reasoning for exclusions being recorded. Disagreements on eligibility were resolved by consensus or through the input of a third investigator. After screening, CitationChaser was used to perform citation chasing on all included studies to identify other potentially relevant studies.^15^ One reviewer [MM, AMS, or DWY] screened the identified papers to decide whether they met the eligibility criteria. Reviewers were not blinded to the journal titles, authors, or institutions.

### Data Extraction and Synthesis

Using a Qualtrics tool, data extraction was conducted by an author [MM, AMS, or DWY] for the following fields for each included paper: article type; article title; publication year; first author; purpose and indication(s) of AI/ML-based medical device; and device users (patients, caregivers, and healthcare providers). Next, the conceptualization and characteristics used to describe barriers and facilitators of transparency and trust and ethical concerns from patients’, caregivers’, and healthcare providers’ perspectives were recorded. For validation, a second reviewer independently performed data extraction on 20% of the final sample, selected at random. Disagreements were resolved by discussion or through the input of a third investigator. Data generated from this project will be actively preserved for three years per Yale Research Data and Materials Policy - Retention 6001.2 unless otherwise required by the journal. Content analyses were performed by MM, using Qualtrics 2022 and Microsoft Excel 2018 (Microsoft Corp) to facilitate data management and organization. We used qualitative coding to categorize the extracted data and identify the key trust and ethical concerns and facilitators.

## RESULTS

### Search Results

The search resulted in 10,171 papers, of which 7,925 were unique. After conducting the first level of screening, 7,799 titles and abstracts were excluded, leaving 126 full-text articles for review. Of those, 71 did not meet eligibility criteria due to ineligible area of care, i.e., non-cardiovascular (n=10); ineligible intervention, i.e., non-AI-ML tools (n=26); ineligible outcome (n=22); ineligible format, i.e., conference abstracts, book chapters, or preprints (n=13), leaving a total of 55 eligible publications. Citation chasing of these articles resulted in 3,603 additional citations, 3,330 of which were eliminated upon title and abstract reviewing. Of the 273 reviewed full-texts, 90 articles were found to be eligible. The reasons for excluding the remaining papers included: ineligible area of care (n=69), intervention (n=14), outcome (n=88), and format (n=12). Overall, 145 papers were included in this review (Figure 1). Since we reached information saturation upon reviewing the additional papers identified through citation chasing, we stopped subsequent rounds of citation chasing.

**Figure 1.**
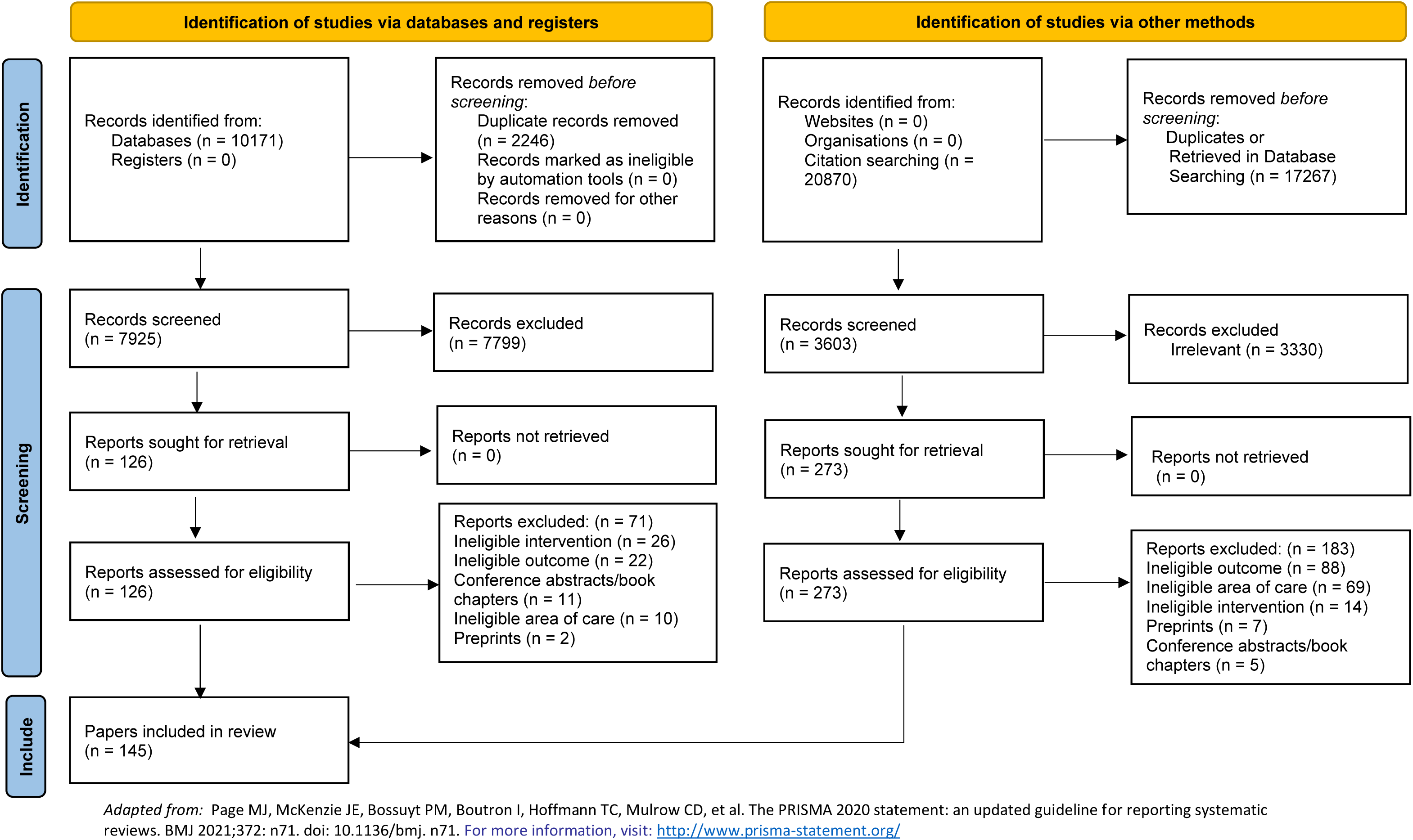
PRISMA Diagram.

### Sample Characteristics

Included articles were published from 2014 to 2022, except for one paper^16^ published in 1996. Of the 145 articles, 88 (60.7%) were review articles; 32 (22.1%) were commentaries, editorials, or perspective pieces; 22 (15.2%) were original research; and 3 (2.1%) were case studies.

The AI/ML-based interventions discussed in 43 (29.7%) papers were devices used for the diagnosis or monitoring of cardiovascular diseases (e.g., AI-enabled cardiac imaging), while 5 (3.4%) were therapeutic devices (e.g., clinical decision support tools for heart pump implants). The interventions discussed in the remaining papers (101 [69.7%]) included both diagnostic and therapeutic AI-based medical devices. The indications for use of the AI-based devices were not specified in most papers (122 [84.1%]). Among those that specified, arrhythmia was the highest reported indication (8 [5.5%]), followed by heart failure (7 [4.8%]). Although all papers discussed AI-based devices in the cardiovascular context, 88 (60.7%) were specific to the cardiovascular specialty, while the remaining articles also included other areas of medicine.

Among all the reviewed articles, 3 (2.1%) studied devices that were self-management software used directly by patients,^17–19^ whereas the main users of the other devices discussed by 48 (33.1%) papers were healthcare providers. The remaining 94 (64.8%) papers did not specify the users. Only 2 (1.4%) papers specified the device sponsor; both studied HeartMan, a personal decision support system for heart failure management, funded by the Horizon 2020 Framework Programme of the European Union.^17,18^

### Ethical Concerns and Mitigation Strategies

There were six key ethical concerns discussed in the literature, which were privacy, security, or confidentiality issues; risk of healthcare inequity or disparity; risk of patient harm; accountability and responsibility concerns; problematic informed consent and potential loss of patient autonomy; and issues related to data ownership (Figure 2). Three papers discussed the lack of human involvement in patient care and the altered relationship between patients and healthcare providers as an ethical concern associated with AI-enabled medical care.^20–22^ One paper debated the additional complexity that AI-based medical devices could add to end-of-life care.^23^

**Figure 2.**
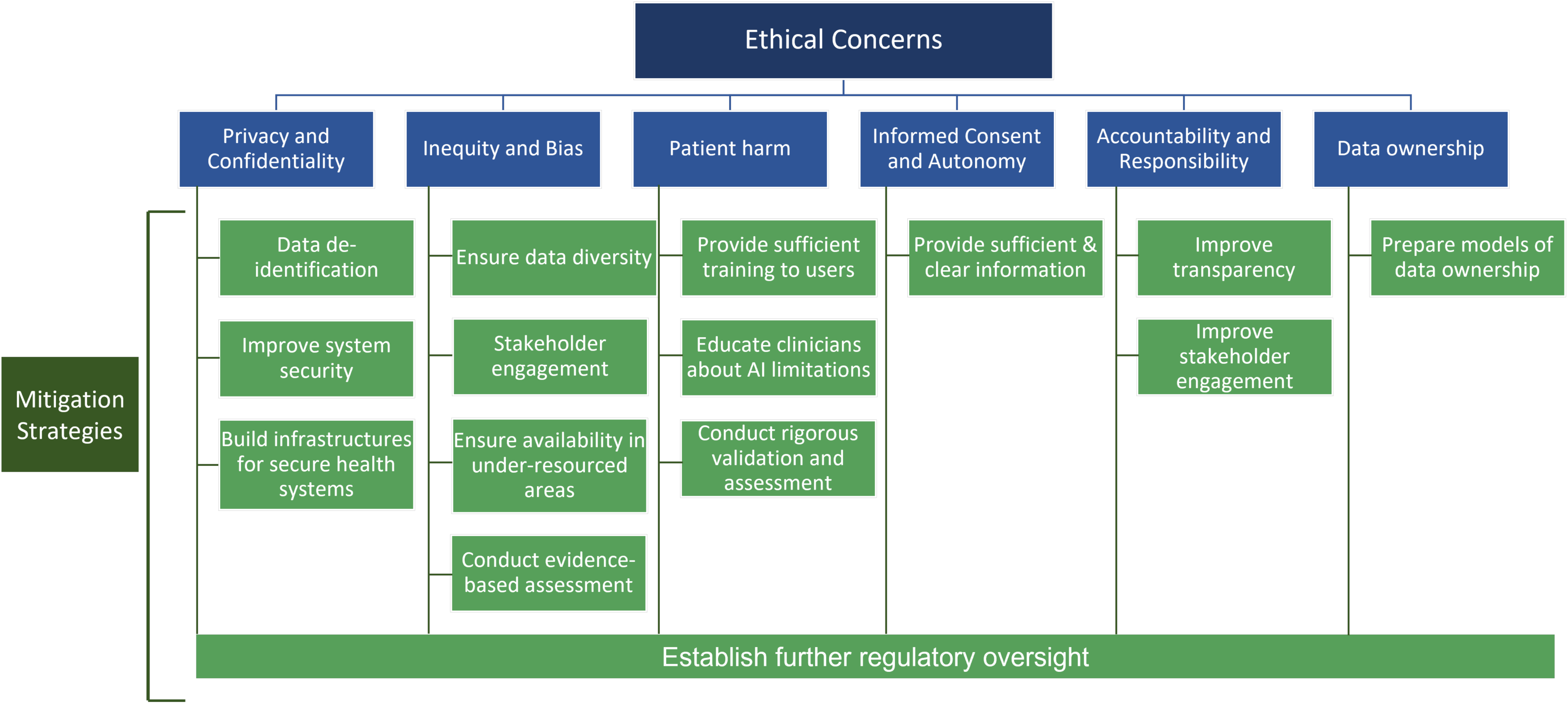
Ethical Concerns and Mitigation Strategies for the Use of AI/ML-based Medical Devices in Cardiovascular Care.

### Privacy, Security, and Confidentiality Concerns

Fifty-nine (40.7%) publications discussed ethical concerns related to privacy, security, or confidentiality. Specific concerns included potential inappropriate access to and misuse of personal information stored in medical devices and inadvertent release of private patient healthcare data.^20,24^ Protecting sensitive patient information from data leakage and cyberattacks, especially for data used by private for-profit organizations,^25^ and protecting the stored medical data, particularly by cloud-assisted AI medical devices or commercial smartphone-based applications with poorly secured servers, were other areas of concern.^26,27^ Moreover, transferring data between institutions for the reproducibility of results could cause additional security problems.^28^ Lastly, ensuring confidentiality could be difficult owing to the circulation of sensitive patient information among unregulated companies and a lack of de-identification of raw data input for AI algorithms.^28,29^

### Mitigation Strategies

We identified mitigation strategies from the literature to address some of the aforementioned ethical concerns. Data de-identification or anonymization and using highly secure data platforms could protect patient data used for the development and training of AI-medical devices.^29–31^ Additionally, more secure health systems across different localities need to be built, and policymakers could help with constructing the adapted infrastructures and developing guidelines regarding patient privacy, data storage, and data sharing to ensure optimal implementation of AI tools in healthcare.^32–34^ Several papers emphasized the need for more regulation and legislation on patient data use, such as performing regular privacy audits, mandating security breach notifications, and setting greater penalties for data misuse.^25,31,35–37^

### Risk of Healthcare Inequity or Disparity

Thirty-six (24.8%) papers raised concerns that AI-based medical devices could create new or exacerbate healthcare inequities or disparities based on factors such as sex, race, ethnicity, or pathology-driven specificities. Potential unfairness in algorithmically automated decisions was described as the major cause of inequities and disparities. Papers discussed the risk of the AI intervention being less effective or providing inaccurate recommendations for under-represented patients if the training datasets for algorithms are based on unrepresentative patient samples.^35,38^ This in turn could lead to discrimination against certain patient populations and increase the gap in healthcare outcomes among different social groups. Furthermore, some were concerned that data could be used to improperly profile patients and differentially provide healthcare (e.g., avoidance of highest-cost or highest-risk patients).^24^ There were also concerns regarding social justice and potential unfairness in the distribution of the benefits and burdens of AI applications.^20^

### Mitigation Strategies

Several papers described important considerations for the data sources used by AI tools to help healthcare providers recognize when it could be inappropriate to use a specific AI tool for certain patient groups and to ensure that access to AI-based tools is not affected by demographic, geographic, or temporal constraints.^39–41^ Strategies to mitigate concerns related to health inequity when using AI in medical care include using a balanced dataset through collecting sufficient data from under-represented populations, validating AI algorithms on different minority and low-income groups, and obtaining robust input from different stakeholders involved in the development, use, and regulation of AI tools.^42–44^ Moreover, creating a distinct algorithm in AI systems for each group of patients, rather than using a universal algorithm for all patients, could improve fairness in decision-making.^45^ Lastly, conducting evidence-based assessment and implementing further regulatory oversights could help to ensure the fairness of AI tools.^26,43^

### Risk of Patient Harm

Concerns about the risk of suboptimal care or patient harm associated with AI tools were raised by 24 (16.6%) papers. Inaccurate data used by AI-based decision tools, flawed AI algorithms, and deliberate hacking of algorithms were discussed as potentially leading to erroneous recommendations and patient harm on a massive scale.^31,46^ The risk of errors would be greater when the AI systems function independently with unchecked decision-making and actions,^47^ particularly in the setting where errors made by complex and untransparent AI systems are difficult to trace and debug.^48^ Moreover, the complexity of AI-based systems, potentially unpredictable system output, and the uncertainty of human–AI interactions could result in substantial variation in the performance of AI-based medical devices, causing further safety challenges.^49^ Lastly, there were concerns about AI-based devices programmed to function in unethical ways, for example by suggesting clinical actions that generate higher profits without patient care benefits.^29^

### Mitigation Strategies

Several papers described the importance of providing sufficient training to device users to reduce the risk of patient harm, with an emphasis on educating healthcare providers about the potential pitfalls and limitations of AI technologies.^46,50^ Additionally, rigorous validation and continuous assessment of the algorithms used in AI-based medical devices, including conducting clinical trials that compare AI-supported care with the standard of care, could identify potential bias in AI algorithms and minimize patient harm.^48,51–53^ Establishing further regulatory and ethical guidelines in the postmarket stage and implementing standard frameworks for regular assessment of the safety of AI tools are also necessary.^31,44^

### Problematic Informed Consent and Loss of Patient Autonomy

We found 17 papers (11.7%) discussing ethical concerns about obtaining informed consent for providing care with AI-enabled medical devices. The main reason leading to problematic informed consent is the lack of transparency and interpretability of AI tools and insufficient information about different aspects of care provided by AI-enabled medical devices.^43,54,55^ Moreover, informing patients about all aspects of health data collection and its use across different platforms and for training algorithms may not be always feasible.^34,56^ Withdrawing consent for the use of these data would cause further challenges.^57^ Eight papers (5.5%) argued that patient autonomy could be negatively affected when using AI-enabled care. This issue specifically is likely to happen if the devices function independently and have unchecked actions,^47^ which could damage patients’ confidence in their ability to change their medical decisions, i.e. refuse care, if later desired.^48^

### Mitigation Strategies

To improve informed decision-making, several papers described the necessity of providing patients and healthcare providers with sufficient information and ensuring that patients are freely able to change their medical decisions if desired.^48,58^ Moreover, further regulations on obtaining valid unambiguous consent when using patient data should be established.^25^

### Accountability and Responsibility Concerns

Another key ethical concern raised by 19 (13.1%) papers was the issue related to accountability and responsibility. Since multiple groups of professionals are involved in the design, manufacture, and use of AI-based medical devices, accountability and liability of the decisions made by these devices could be difficult to determine. While some suggested that users of the devices should ultimately be responsible for the output of algorithms,^23,59^ there are considerable debates around the accountability of actions suggested or performed by AI-based technologies and the potential misuse of data.^34,35^ The complexity, opaqueness, and lack of transparency of AI-based medical devices make the accountability and responsibility issues even more challenging.^48,60^

### Mitigation Strategies

To address questions of accountability, several papers described the importance of improving the engagement of all stakeholders, including physicians and developers. Papers also suggested improving the transparency of AI tools’ function so that the reasons behind decisions and actions taken by the devices are clear.^61,62^ Moreover, there is a need for regulatory and legal systems to oversee the implementation of AI-based medical devices and determine the responsibilities of patients, healthcare providers, and others.^63^

### Data Ownership Issues

There were further ethical concerns discussed by 11 (7.6%) papers related to ownership of the patient data being used by AI-based technologies, particularly if the data is identifiable.^64^ The rules and regulations related to data ownership vary significantly across different regions and may be absent in some jurisdictions, which makes it unclear whether patients, hospitals, or private companies own the data analyzed by AI tools.^65,66^ This issue is directly associated with how AI and its data are monetized,^66^ as there are controversies about who should profit from the collected data and for how long these institutions or individuals can and should retain patient health information.^67^

### Mitigation Strategies

To address these concerns, several papers described the importance of clear regulations around data ownership and preparing models of health data ownership with rights to the individual ahead of using AI-based devices in healthcare. ^31,36^

### Trust Barriers and Facilitators

We identified 53 (36.6%) and 58 (40.0%) papers discussing trust barriers and facilitators, respectively, from patients’ and healthcare providers’ perspectives when using AI-based medical devices in cardiovascular care (Figure 3).

**Figure 3.**
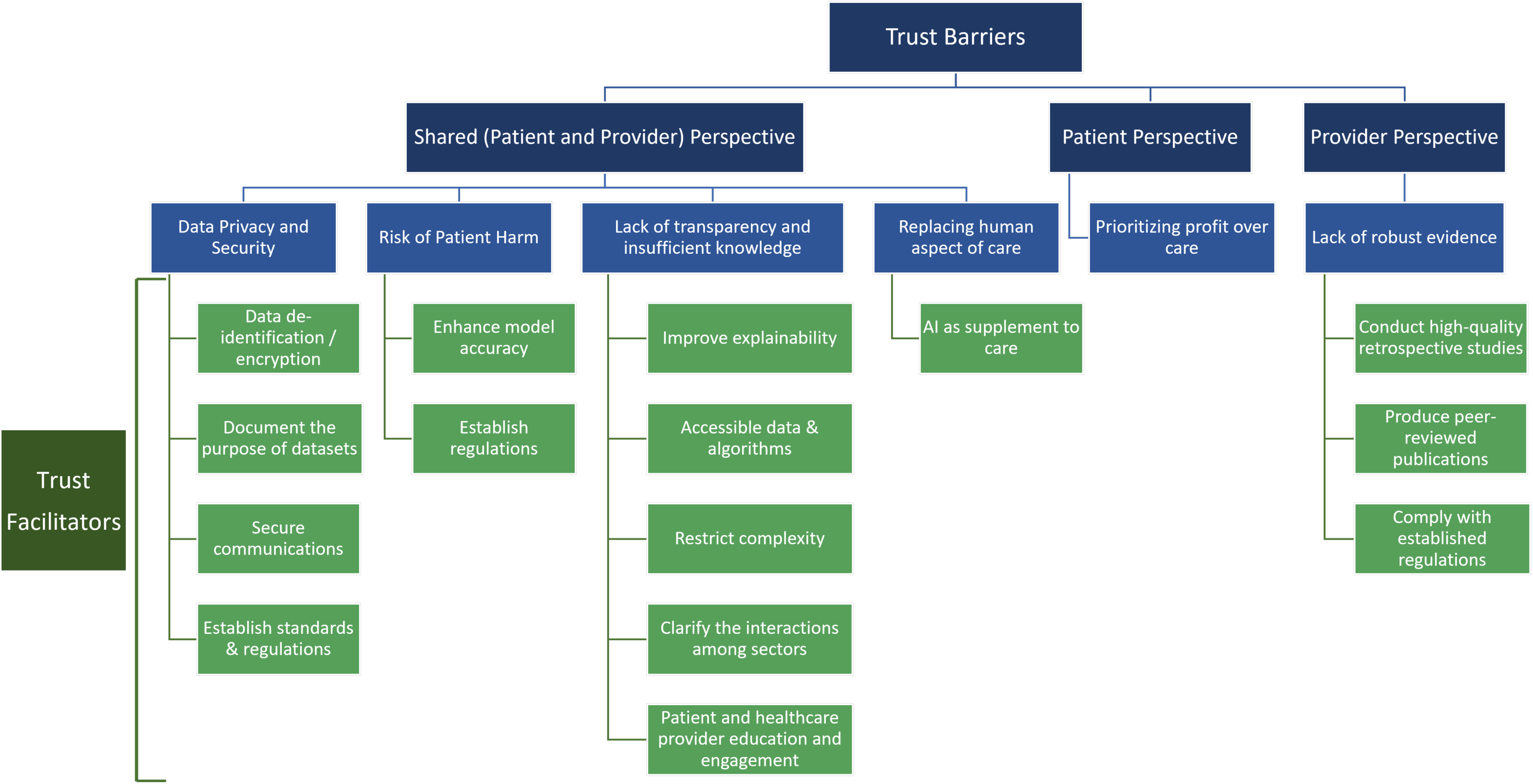
Trust Barriers and Facilitators for the Use of AI/ML-based Medical Devices in Cardiovascular Care.

#### Shared (Patient and Healthcare Provider) Perspective

### Data privacy and security issues

Data privacy and security concerns were discussed as key trust barriers for patients and healthcare providers.^17,62^ In particular, patients were described as worried about the potential alteration of data, unauthorized use of data, information sharing with commercial partners, and data loss.^57,68^ These issues are specifically concerning in the absence of uniform federal privacy regulations regarding collecting, storing, and using patient health information in different settings.^39^

### Facilitators

To address data privacy and security concerns, the literature discussed encrypting patient data according to the Health Insurance Portability and Accountability Act of 1996 (HIPAA), removing data identifiers, documenting the purpose of datasets, establishing ethical standards for data use and access, and securing communications between patients and healthcare providers.^39,69,70^ Regulatory bodies could ensure the competence of AI systems and their users and establish standardized codes of ethics and conduct for device developers.^70^

### Risk of Suboptimal Care or Patient Harm

Users have expressed concerns around the possibility of device malfunction and are hesitant about the trustworthiness of diagnostic decisions or automatically generated medical advice by AI tools, especially if the advice contradicts their previous experiences.^48^ Another important trust barrier is the uncertainty about the reliability and quality of the data used in the algorithms, which could be incomplete, unrepresentative, or outdated.^71^ This lack of generalizability could exacerbate health inequities, and further decrease trust in the populations who feel that AI would be inaccurate when applied to their cases.^72^ Certain populations may also feel that they may not equally benefit from AI technologies because of the deployment and marketing strategies that manufacturers might take.^72^ Healthcare providers are also concerned that AI-based medical devices could provide inaccurate or biased recommendations, especially if the systems are not regularly updated.^73,74^ Moreover, clinicians may not trust the generalizability of the outputs of AI systems for their own patients due to the lack of diversity in the clinical dataset.^75–77^

### Facilitators

To address these trust barriers, the literature discussed the importance of keeping AI systems updated by introducing new rules and cases along with routine performance assessments to enhance the accuracy of decisions made by AI-based medical devices.^73,78^ Further regulations and legislation could also increase trust by ensuring the balance between innovation and patient safety and confirming that AI algorithms meet appropriate standards of clinical benefit.^79,80^

### Lack of transparency and Insufficient knowledge

Substantial barriers to trust in AI-enabled medical devices are the lack of transparency, opaqueness (black box nature), and poor interpretability of the devices.^74,81,82^ Physicians tend to trust a device less if they do not fully understand how it functions or how its outputs are generated, even if the device performs well.^35,38,52^ Multiple barriers to transparent AI-based medical devices exist, including the lack of understanding of what information is being used by the AI tools, what the AI systems are learning, and how the AI algorithms reach conclusions based on the inputs.^28,83–85^ Also, it could be difficult to achieve algorithmic transparency due to the complicated structure, dynamic learning, and constant evolution of AI algorithms.^34,54^ These factors make AI models difficult to explain and justify, and therefore, uninterpretable.^86^ Besides, inadequate education and experience with AI tools can cause additional barriers to trustworthy AI-enabled care.^74,87^

### Facilitators

To improve explainability and physicians’ understanding of AI-based medical devices, it is essential to clarify AI algorithm training data, explain the computational model and its output, and acknowledge the existing limitations of AI-based medical devices.^74,76,85,88,89^ Making the datasets, codes, and trained models publicly available and using interpretable models that will allow healthcare providers to review and provide feedback to the AI decision-making tools could further improve transparency.^45,90^ Some argued that healthcare providers may not need detailed explanations of the validated predictions and decisions made by AI-enabled medical devices but need to have sufficient information about the major components that affect the decisions.^41^ Additionally, a visual display of the consensus between decision support tools and clinicians’ assessments could enhance clinicians’ trust in AI systems.^53^

Restricting the complexity of AI tools as well as providing clarity on how AI devices are regulated could facilitate patient trust.^17,19,57,91^ It is also essential to provide patients with appropriate education about how to use AI tools and enhance their engagement in different phases of the design and implementation of AI technologies.^48,87,92,93^

Other important factors for facilitating transparency are to clarify all the interactions within and among different sectors that led to the development of AI systems and to maintain open and clear communication between healthcare providers and developers.^86,94^ Regulatory bodies could establish more rigorous regulations for the enforcement of transparency in datasets and algorithms used in AI-based medical devices.^45,90^

#### Replacing human aspects of care

Patients and healthcare providers seem to trust AI tools less if the devices are meant to entirely replace the human aspect of care.^51^

### Facilitators

Trust could improve if patients and healthcare providers are assured that AI-based devices are supplementary to care, rather than outright replacing clinicians or other human aspects of care.^51,90^

#### Patient Perspective

### Prioritizing profits over patients’ interests

From the patient perspective, trust would be diminished when they feel AI devices are mainly used for economic efficiency at the cost of patient interests and benefits.^70^

No facilitators were identified in the reviewed literature for this trust barrier.

#### Healthcare Provider Perspective

##### Lack of robust evidence

A significant barrier to clinician trust is the lack of robust evidence for the accuracy and limitations of AI-based medical devices in addition to the inadequate education and training about the use of AI tools.^74,95,96^

### Facilitators

Several papers argued that while it might not be feasible to explain all aspects of AI, generating more reliable evidence and standards through rigorous internal and external validations, prospective clinical trials in diverse cohorts which demonstrate safety, efficacy, and generalizability of AI devices, and peer-reviewed publications can improve trust.^97–101^ Therefore, collaborative practices with healthcare providers for the development and continuous assessment of AI devices are essential.^73,96^ Lastly, complying with the established legislations and regulations is essential when producing trustworthy AI research.^86^

## DISCUSSION

After reviewing more than 11,000 publications, we identified key ethical concerns and major trust barriers around the use of AI in cardiovascular care held by healthcare providers and patients. Concerns focused on data privacy and security, risk of patient harm, and the possibility that AI/ML-based medical care could exacerbate healthcare inequities or advance unfair algorithmically automated decisions. Inadequately obtaining informed consent from patients regarding the use of AI and various forms of data collection while providing AI-enabled care was also described, as was determining who is ultimately responsible for regulating the development, performance, and use of AI in medicine and who owns the collected data. The absence of rigorous clinical trials to support the safety and efficacy of AI-enabled medical devices and the lack of transparency about the data used by AI devices and their subsequent recommendations remain other significant barriers to patients’ and healthcare providers’ trust. These challenges should be carefully identified and addressed to ensure that AI systems are developed and implemented in an ethical and trustworthy manner.

We identified mitigation strategies to address most key ethical and trust concerns about the use of AI in medicine, which requires a collaborative effort involving AI developers, regulators, hospital systems, healthcare providers, and patients. Regulatory agencies were identified as having multiple inroads to addressing patient and clinician concerns. Notably, we found that establishing further regulations and legislation around development, adoption, and use of AI in healthcare is a key facilitator for addressing almost all the identified ethics concerns and trust barriers. Certain proposed frameworks and guidance documents have carved out actions for oversight bodies to delineate the scope of liability, strengthen data privacy protections, and clarify data ownership regulations.^102,103^ Moreover, requiring postapproval studies could ensure continuous monitoring of AI devices’ performance, potential biases, and unintended consequences.

AI developers similarly have a significant stake in addressing patient and clinician concerns and need to be attentive to data stewardship practices, safety, and transparency as models are researched, developed, and marketed. Moreover, current medical device labeling does not always address the unique challenges of the use of AI/ML-based software, such as training data sources, model accuracy, potential biases, and opting out of use, which can hinder patient-shared decision-making and trust in AI-enabled care. Providing AI model facts labels will establish a clear and standardized communication of information with users and enhance transparency and trust.^50^ Furthermore, self-governance approaches may serve as a potential mechanism in tandem with regulatory intervention for implementing mitigation strategies. Submitting to a set of industry standards as well as certification processes may help to mitigate the risks of AI tools and help to facilitate trust in models.^104^

Hospital systems and clinicians will also be faced with key decisions regarding AI tools adopted in their practices. As hospitals become a source of data for the development of numerous models, appropriate privacy protections and transparency about data use and model deployment would be relevant, especially as they act in coordination with third-party developers.^105^ As end-users of most healthcare AI tools, clinicians may become responsible for providing appropriate information about these systems to patients at the point of care and for appropriately integrating model insights into clinical decision-making.

While our findings are indicative of many strategies that would be taken up by clinical, technical, and regulatory stakeholders, there are also opportunities for including patients. Stakeholder engagement with patient populations and the public in the research and design of AI tools may be relevant to mitigating bias and developing trust, particularly by communicating the underlying design of AI tools in ways that are understandable to patients and leveraging advisory groups to inform the creation of such tools.^106^ Identifying opportunities for patient engagement will be incumbent upon all stakeholders with more formal decision-making authority. Thus, regulatory oversight on using and sharing patient information, safety and transparency of AI tools, and responsibilities of healthcare providers, device manufacturers, and patients would facilitate the application of AI in medical care.

Overall, we found that most papers briefly touched upon issues related to trust and ethics and potential mitigation strategies without providing in-depth information. Additional studies translating ethical principles into tangible tools and guidance for stakeholders will be an important next step in implementation of responsible and trustworthy AI-enabled health care.^107^ Moreover, we did not find any ethical concerns or trust barriers and facilitators from the caregivers’ perspective, necessitating further research in this area.

Our study has limitations. First, similar to all reviews of published literature, publication and reporting biases may have affected our findings. Second, while we identified and reviewed a significant number of relevant papers, the vast majority were review articles and commentaries, editorials, or perspective pieces with fewer original research articles. While our search was very exhaustive, there was an inconsistency in the level of detail, which may have led to papers potentially being missed. However, citation chasing was undertaken to identify additional relevant articles that failed to include the three main concepts of our search. Lastly, this study focused on the use of AI in cardiovascular care and may not generalize to uses in other areas of medicine.

## CONCLUSION

This literature review of using AI/ML-based interventions in cardiovascular care identified key ethical and trust concerns from patients’ and healthcare providers’ perspectives, including issues related to data privacy and security, potential inequity and bias, risk of patient harm, patient consent and autonomy, and a lack of transparency about the function of AI-based medical devices. To address these concerns, certain mitigation strategies, particularly establishing further regulatory oversight on the use of patient data, and safety and transparency of AI tools seem necessary.

## List of abbreviations

AI: Artificial Intelligence
HIPAA: Health Insurance Portability and Accountability Act of 1996
ML: Machine learning

## Declarations

### Ethics approval and consent to participate

Ethical approval was not required. Publicly available nonclinical datasets were used. Informed consent was not needed because no patient data were used.

### Consent for publication

Not applicable.

### Availability of data and materials

Relevant data are available on reasonable request from the corresponding author.

### Authors’ contributions

JM and MM are the guarantors of the review. MM and JM drafted the protocol. MM, JM, and AAG developed the search strategy with input from all the authors. MM, AAG, AMS, and DWY screened the articles and extracted the findings. MM summarized the data and wrote the first draft of the article. All authors critically reviewed and revised the manuscript for publication.

## Data Availability

Relevant data are available on reasonable request from the corresponding author.

## Acknowledgments

Not applicable

