## Appendix 1 for "Barriers and Facilitators to Trustworthy and Ethical AI-enabled Medical Care From Patient’s and Healthcare Provider’s Perspectives: A Literature Review"

**Ovid Embase**

1	(exp cardiovascular disease/ or exp cardiac equipment/) and (exp artificial intelligence/ or exp machine learning/)

2	(heart or cardiac or myocard* or arrhythmia* or valvular or coronar* or cardio* or ventric* or hypertens* or vascular or angiogra* or echocardiogra* or electrocardiogra* or EKG or ECG or pacemaker*).tw,kf.

3	((artificial or computat* or computer* or machine or deep or transfer or hierarchical) adj3 (intelligence* or learning* or reasoning* or diagnosis or diagnoses or simulation* or interface* or assist*)).tw,kf.

4	(neural network* or random forest* or decision tree* or knowledge representation* or computer vision system* or computer reasoning*).tw,kf.

5	((predict* or prognost* or clinical decision) adj3 (support* or system* or model* or tool* or scoring or calculation* or algorithm*)).tw,kf.

6	AI.ti,ab.

7	3 or 4 or 5 or 6

8	2 and 7

9	1 or 8

10	exp patient attitude/

11	(exp trust/ or exp knowledge/ or exp perception/ or public opinion/ or uncertainty/) and exp patient/	40190

12	((patient* or consumer* or care-giver* or caregiver*) adj3 (accept* or adherence or attitude* or awareness* or barrier* or behavior* or behaviour* or belief* or choice* or compliance or confidence or decision* or dislik* or distrust* or doubt* or facilitator* or fear* or hesita* or knowledge or mistrust* or motivat* or nonadherence or non-adherence or noncompliance or non-compliance or opinion* or perception* or preference* or reluctan* or refus* or trust* or understand* or understood or uptak* or willingness)).tw,kf.

13	10 or 11 or 12

14	9 and 13

**Ovid MEDLINE(R) ALL**

1	exp cardiovascular diseases/ and (exp artificial intelligence/ or exp machine learning/)

2	(heart or cardiac or myocard* or arrhythmia* or valvular or coronar* or cardio* or ventric* or hypertens* or vascular or angiograph*).tw,kf.

3	((artificial or computat* or computer* or machine or deep or transfer or hierarchical) adj3 (intelligence* or learning* or reasoning* or diagnosis or diagnoses or simulation* or interface* or assist*)).tw,kf.

4	(neural network* or random forest* or decision tree* or knowledge representation* or computer vision system* or computer reasoning*).tw,kf.

5	((predict* or prognost* or clinical decision) adj3 (support* or system* or model* or tool* or scoring or calculation* or algorithm*)).tw,kf.

6	3 or 4 or 5

7	2 and 6

8	1 or 7

9	exp Health Knowledge, Attitudes, Practice/

10	exp Attitude to Health/

11	(exp trust/ or exp knowledge/ or exp perception/ or public opinion/ or uncertainty/) and exp patient/

12	((patient or consumer or care-giver* or caregiver*) adj3 (accept* or adherence or attitude* or awareness* or barrier* or behavior* or behaviour* or belief* or choice* or compliance or confidence or decision* or dislik* or distrust* or doubt* or facilitator* or fear* or hesita* or knowledge or mistrust* or motivat* or nonadherence or non-adherence or noncompliance or non-compliance or opinion* or perception* or preference* or reluctan* or refus* or trust* or understand* or understood or uptak* or willingness)).tw,kf.

13	or/9-12

14	8 and 13

**Scopus**

( TITLE-ABS-KEY ( ( patient* OR consumer* OR care-giver* OR caregiver* ) W/3 ( accept* OR adherence OR attitude* OR awareness* OR barrier* OR behavior* OR behaviour* OR belief* OR choice* OR compliance OR confidence OR decision* OR dislik* OR distrust* OR doubt* OR facilitator* OR fear* OR hesita* OR knowledge OR mistrust* OR motivat* OR nonadherence OR non-adherence OR noncompliance OR non-compliance OR opinion* OR perception* OR preference* OR reluctan* OR refus* OR trust* OR understand* OR understood OR uptak* OR willingness ) ) ) AND ( TITLE-ABS-KEY ( ( artificial OR computat* OR computer* OR machine OR deep OR transfer OR hierarchical ) W/3 ( intelligence* OR learning* OR reasoning* OR diagnosis OR diagnoses OR simulation* OR interface* OR assist* ) ) OR TITLE-ABS-KEY ( "neural network*" OR "random forest*" OR "decision tree*" OR "knowledge representation*" OR "computer vision system*" OR "computer reasoning*" ) ) AND ( TITLE-ABS-KEY ( heart OR cardiac OR myocard* OR arrhythmia* OR valvular OR coronar* OR cardio* OR ventric* OR hypertens* OR vascular OR angiogra* OR echocardiogra* OR electrocardiogra* OR ekg OR ecg OR pacemaker* ) )

**Web of Science Core Collection**#1 TS=(heart or cardiac or myocard* or arrhythmia* or valvular or coronar* or cardio* or ventric* or hypertens* or vascular or angiogra* or echocardiogra* or electrocardiogra* or EKG or ECG or pacemaker*)

#2 **TS=((artificial or computat* or computer* or machine or deep or transfer or hierarchical) near/3 (intelligence* or learning* or reasoning* or diagnosis or diagnoses or simulation* or interface* or assist*)) or TS=("neural network*" or "random forest*" or "decision tree*" or "knowledge representation*" or "computer vision system*" or "computer reasoning*") or TS=**

**((predict* or prognost* or "clinical decision") near/3 (support* or system* or model* or tool* or scoring or calculation* or algorithm*))**

**#3 TS=((patient* or consumer* or care-giver* or caregiver*) near/3 (accept* or adherence or attitude* or awareness* or barrier* or behavior* or behaviour* or belief* or choice* or compliance or confidence or decision* or dislik* or distrust* or doubt* or facilitator* or fear* or hesita* or knowledge or mistrust* or motivat* or nonadherence or non-adherence or noncompliance or non-compliance or opinion* or perception* or preference* or reluctan* or refus* or trust* or understand* or understood or uptak* or willingness))**

**#4 #1 and #2 and #3**

**Cochrane Library**

#1 ((patient* or consumer* or care-giver* or caregiver*) near/3 (accept* or adherence or attitude* or awareness* or barrier* or behavior* or behaviour* or belief* or choice* or compliance or confidence or decision* or dislik* or distrust* or doubt* or facilitator* or fear* or hesita* or knowledge or mistrust* or motivat* or nonadherence or non-adherence or noncompliance or non-compliance or opinion* or perception* or preference* or reluctan* or refus* or trust* or understand* or understood or uptak* or willingness)):ti,ab

#2 ((artificial or computat* or computer* or machine or deep or transfer or hierarchical) near/3 (intelligence* or learning* or reasoning* or diagnosis or diagnoses or simulation* or interface* or assist*)):ti,ab or ("neural network*" or "random forest*" or "decision tree*" or "knowledge representation*" or "computer vision system*" or "computer reasoning*"):ti,ab or

((predict* or prognost* or "clinical decision") near/3 (support* or system* or model* or tool* or scoring or calculation* or algorithm*)):ti,ab

#3 (heart or cardiac or myocard* or arrhythmia* or valvular or coronar* or cardio* or ventric* or hypertens* or vascular or angiogra* or echocardiogra* or electrocardiogra* or EKG or ECG or pacemaker*):ti,ab

#4 #1 and #2 and #3

**Google Scholar**

Artificial intelligence cardiovascular trust
